# Analysis of psychological state and clinical psychological intervention model of patients with COVID-19

**DOI:** 10.1101/2020.03.22.20040899

**Authors:** Lu Yang, Danjuan Wu, Yanbin Hou, Xunqiang Wang, Ni Dai, Guanjun Wang, Qing Yang, Wenhui Zhao, Zhongze Lou, Yunxin Ji, Liemin Ruan

## Abstract

**Background:** Patients with the 2019 coronavirus disease (COVID-19) have different degrees of psychological pain, such as anxiety and depression, which may related to their prognosis. Psychological intervention can be conducted in different ways to improve psychological pain and improve the treatment effect.

**Objective:** The present study aimed to investigate and analyze the psychological status of patients with COVID-19 during the course of illness, and to evaluate the effect and influencing factors of psychological crisis intervention, so as to explore the effective mode of clinical psychological intervention in acute patients under isolation environment.

**Methods:** A total of 143 persons participated in the study, including 26 patients diagnosed with COVID-19 in the isolation ward (COVID-19 group), 87 patients with general pneumonia in the observation ward (General Pneumonia group) and 30 healthy volunteers (Normal group). All the patients in the ward received comprehensive psychological intervention, including telephone psychological counseling (active and passive), self-adjustment of written materials and one-to-one psychological crisis intervention. Hamilton depression scale (HAMD) and Hamilton anxiety scale (HAMA) were used to evaluate the mental health status of all patients on the day of admission and 1 week after treatment.

**Results:** The scores of HAMA and HAMD of all patients (including isolation ward and observation ward) were significantly higher than the healthy volunteers at the time of admission. The total score of HAMA and HAMD in CVOID-19 group were both higher than that General Pneumonia group. After 1 week’s hospitalization with comprehensive psychological intervention, the scores of HAMA and HAMD in CVOID-19 group were significantly decreased.

**Conclusion:** Patients those who diagnosed with COVID-19 in the isolation ward and/or general pneumonia in observation ward have different degrees of anxiety, depression and sleep problems. While receiving antiviral treatment, patients also need psychological intervention. Comprehensive psychological intervention model has been proved to be effective.

## INTRODUCTION

The 2019 coronavirus disease (COVID-19) is a new type of respiratory infections, since December, 2019, in Wuhan, Hubei Province, China, which has spread rapidly throughout the country, and has now spread to the global infectious diseases, this is strong, high-rise close contact with the outbreak of the epidemic. A few cases were serious or even fatal ^[1-3]^. The rapid spread of the disease in a wide range of vulnerable to varying degrees of panic, not conducive to the disease for treatment and rehabilitation. Ningbo First hospital as a designated hospitals for COVID-19 admitted that the disease medical care for patients in a timely and effective psychological intervention is also an important task. As pneumonia patients need strict isolation, medical staff need to strictly guard, combined with mobile phones, networks, and the establishment of the psychological advisory group^[4]^, a phone, text, video and one-to-one online psychological intervention and so on, in order to explore the hospital confirmed and suspected new corona virus in the disease process, the patient’s psychological changes, as well as psychological intervention effectiveness.

## MATERIAL AND METHODS

### Participants and study design

Collection 2020 February 5 to February 29 in view of the patient wards, including a total of 143 cases of 26 patients diagnosed with COVID-19 (COVID-19 group), who are in line with the interim guide diagnostic standards and national defense construction committee of the novel coronavirus pneumonia clinical trial program (version 6), diagnostic criteria, and nasopharyngeal swab specimen by Nucleic Acid Detection tests positive, of which 9 were men (35 per cent), 17 were women (65 per cent), and the age up to 86 years of age, minimum age of 27 years old, average age 56, mostly elderly patients. General Pneumonia group: 119 patients with general pneumonia met the diagnostic criteria of pneumonia, but the Nucleic Acid Detection test of the pharyngeal swab was negative, and 32 patients with general pneumonia were excluded due to unwillingness to participate or other reasons can’t complete the study (including hearing impairment, speech impairment).

There are 114 patients (COVID-19 group n=26 and General Pneumonia group n=87) with comprehensive psychosocial interventions, including telephone counselling (active and passive), written materials self-adjustment and one-to-one psychological crisis intervention. Normal Group: As the age and sex of the patient to match the healthy volunteers, a total of 30, of which 10 were male (33%), 20 (67%), with an average age of 52 years.

This study was approved by the Ethical Committee of Ningbo First Hospital (approval number: 2020-R042) and registered with the registry website *http://www.chictr.org* (registration number: ChiCTR2000030697). All participants in this study signed informed consent was obtained.

The route of present study as shown in the figure1.

**Figure 1:**
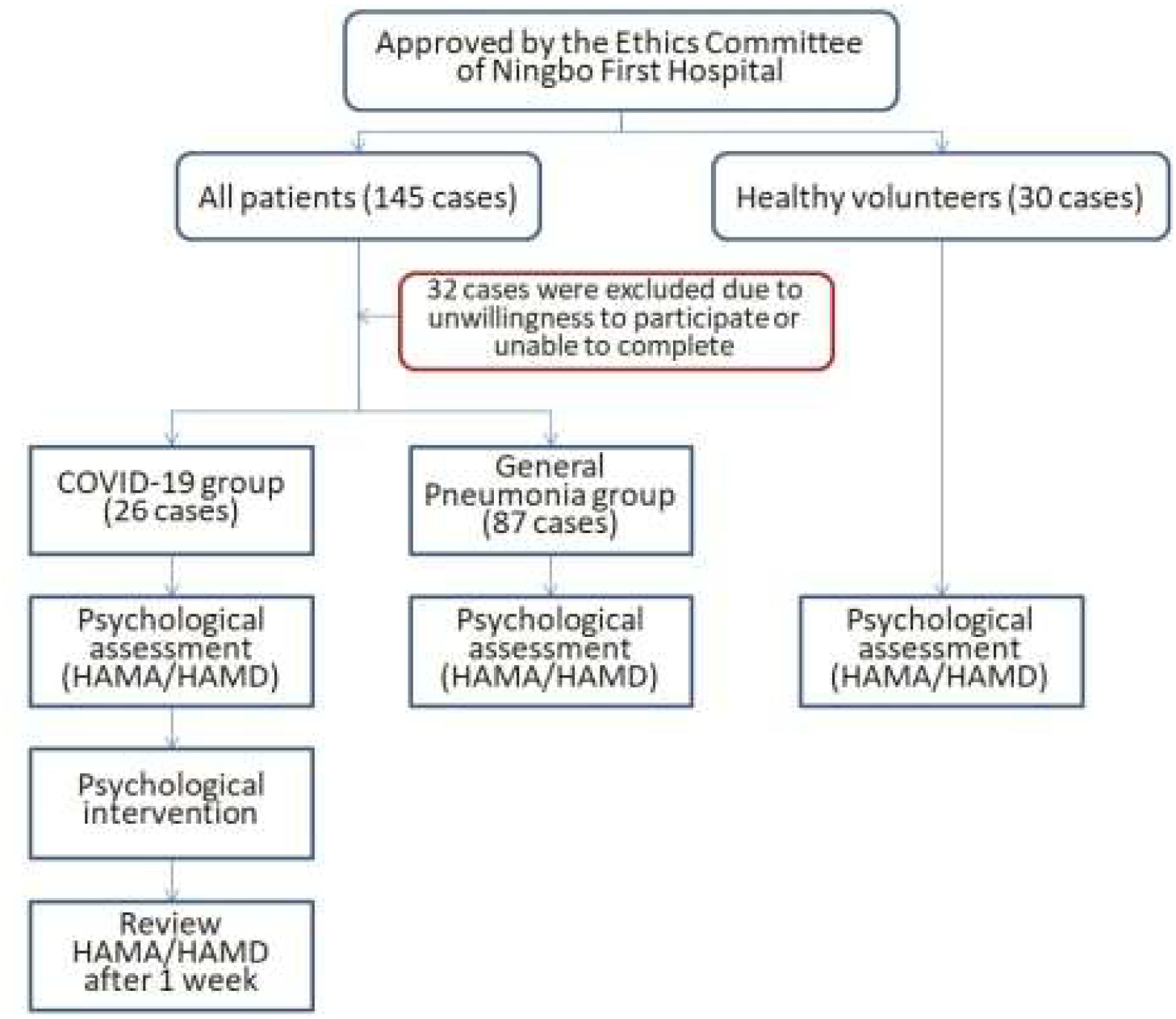
Design route of this study.

### Data and Instruments

Participants provided their sociodemographic information and clinical illness histories and completed several measures of psychosocial functioning at study baseline. Using the Hamilton depression scale (24) (A HAMD-24) and Hamilton anxiety gauge (HAMA) assess all patients and healthy cases and integrated psychological intervention, including telephone counselling (active and passive), written materials self-adjustment and one-to-one psychological intervention. After 1 week, the diagnosis for all coronavirus pneumonia patient re-evaluated.

The main statistical indicators of HAMD-24 score and HAMA total score, HAMD-24 total score is less than 8 points and there is no depression, 8-20 points may have depression, 20-35 points certainly depression, 35 points or more severe depression; HAMA total score is less than 7 points without anxiety, 7-14 points may have anxiety, 14-21 points certainly anxiety, 21-29 points must be a clear anxiety, 29 points or more serious anxiety.

### Statistical analysis

Statistical survey database established by using social science software SPSS19.0, and statistical analysis, by analysis of variance for multiple samples of mean difference comparison, by t test for paired samples of mean differences, p < 0.05 for the difference was statistically significant.

## RESULTS

### Analysis of psychological state with coronavirus pneumonia on admission

The scores of HAMA and HAMD-24 in patients with COVID-19 and general pneumonia were significantly higher than the healthy volunteers at the time of admission. The results as shown in table 1.

**Table 1:** The scores of HAMA and HAMD-24 in COVID-19 group and General pneumonia group compared with the Normal group.

|  | CVOID-19 group | General Pneumonia group | Normal group | p-value |
| --- | --- | --- | --- | --- |
| HAMD-24 total score | 5.96±3.34 | 2.77±2.73 | 1.47±1.41 | <0.005 |
| HAMA total score | 7.85±2.56 | 4.29±3.62 | 1.60±1.63 | <0.005 |

### The effects of psychological intervention on COVID-19 patients

At the time of admission, the anxiety and depression of patients with COVID-19 was obviously. Through 1-week hospitalization with psychological intervention and daily telephone contact for supportive psychological intervention, the scores of HAMA and HAMD-24 in CVOID-19 group were significantly decreased. As shown in figure 2.

**Figure 2:**
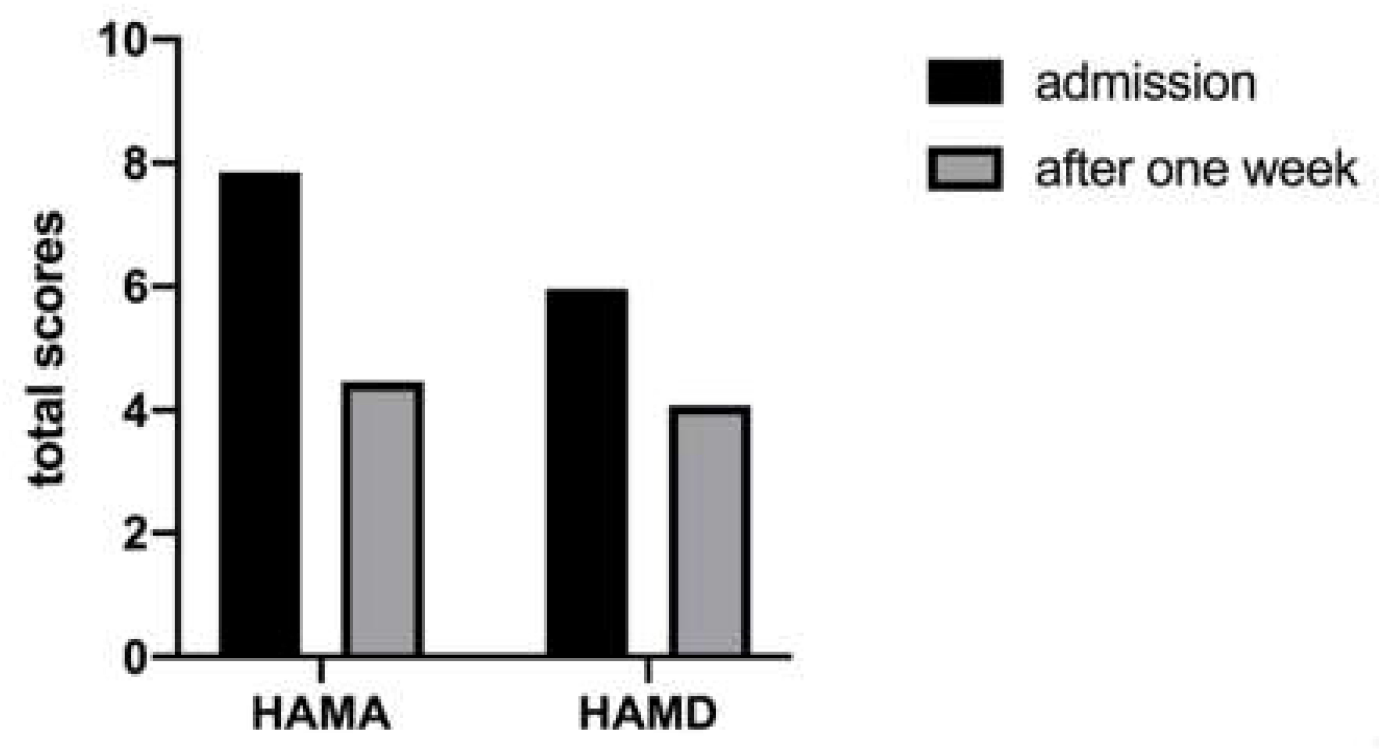
The effects of psychological intervention on COVID-19 patients. Through 1-week hospitalization with psychological intervention and daily telephone contact for supportive psychological intervention, the scores of HAMA and HAMD-24 in CVOID-19 group were significantly decreased.

## DISCUSSION

The main findings of the present study were that HAMA total score of hospitalized patients were generally high. That is common in hospitalized patients with varying degrees of psychological problems, especially have a higher level of anxiety and somatization symptoms[4], suggesting that patients not only bear the severe physical pain, and mental distress, especially the COVID-19 patients with anxiety depression is more common in patients with general pneumonia, consider the reason has the following several aspects: 1. From the perspective of the route of transmission of COVID-19, mainly for the close contact and the spread of respiratory tract, the infectious, many for family gathering, patients not only concerned about their illness, more concerned about the infection of members of the family; 2. From the perspective of COVID-19, the new virus had never found in the human body, because of Wuhan viral pneumonia cases were found in 2019, was named the world health organization (WHO) on January 12, 2020, so the people the knowledge of the new disease, less about the characteristics of the virus and disease diagnosis, treatment and prognosis are exploring, so the uncertainty is adding to the patients with tension and fear of disease; 3. In a short period of time, a large number of COVID-19 patients broke out, the news media continuously reported, the death of some patients, all make the patients’ psychological bearing capacity is low, in the course of illness is prone to depression, or even negative ideas. Therefore, in the process of disease treatment, patients not only need medical treatment, but also need psychological support. It is worth noting that in the process of receiving the intervention, a small number of patients began to reject the psychological intervention and deny the psychological problems. After our patient and careful explanation and communication, they finally accepted our help, which also reflected that our medical staff needed to take the initiative to publicize and provide help to more patients.

COVID-19 is a strong contagious acute respiratory disease, the course of development is fast, the symptom such as patients with high fever, difficulty breathing[1-3,5], there is no specific medicine, treatment is more suit the support is given priority to, appear easily in patients with anxiety and panic, and hospitalization after strict isolation environment cannot receive relatives and friends of emotional support, message block, prone to pessimism and despair, often have a lot of psychological pressure, was in a state of crisis, the urgent need for psychological support and intervention. But as a result of the new virus infectivity, mainly for the droplet and contact transmission in close communication, the traditional face-to-face psychological crisis intervention, on the one hand, easy to cause disease is on the other hand also increases medical supplies such as isolation of a large number of consumption, are more likely to cause cross infection, and telephone psychological crisis intervention is effective to avoid the spread of the virus, but also has better timeliness. Our daily morning patients by phone or radio link, understand the patients psychological condition at present, the problems existing in the process of its life or treatment timely communication and solve, stable patients with their own emotions, and disease education, more positive information feedback to the patient, strengthen the positive emotions, increase treatment adherence, cooperate to ward treatment, prevent impulse to run, self-injury, hurt, such as abnormal behavior, adverse effects on individuals, families and society.

This study has some limitations, as a result of the new champions league pneumonia in patients with less sample size, research results have representative need to further expand the study sample size, and the patient’s isolation and protection of medical personnel to carry out psychological intervention also caused a certain limit, psychological treatment in addition to the speech communication face to face, and even more expressions and gestures, and medical personnel’s body language also have obvious effect on patients with stable mood, such as watching a nod, take back, thumb, V gestures, etc., can better close distance, make patients feel be respected and valued, psychological assessment and psychological intervention on the patients better, promote disease rehabilitation. In addition, the psychological intervention without using case-control study, it is difficult to eliminate the effects of disease rehabilitation of mental state at the same time the author also noted that the COVID-19 outbreak is long-term and far-reaching impact on patients, as the patients’ rehabilitation home may appear new problems such as post-traumatic stress disorder, at present the related content need further follow-up, relevant data also in the collecting and organizing of further, we will further complete the next stage.

## CONCLUSIONS

Understanding the psychological state of patients with pneumonia, especially patients with COVID-19, can help clinicians to systematically identify patients vulnerable to psychological pain, and provide targeted psychosocial interventions to improve the mental health of patients.

## Data Availability

The datasets generated during and/or analysed during the current study are available from the corresponding author on reasonable request,
the partial datasets generated or analysed during the current study are included in this preprint article and not publicly available due to formally published, but are available from the corresponding author on reasonable request.

## AUTHORS’ CONTRIBUTIONS

LY and ZZL drafted the manuscript, LMR, YXJ and ZZL conceived and designed framework of this article, LY, YBH, XQW, ND, GJW, QY WHZ and YXJ collected and analyzed the literature. All authors read and approved the final manuscript.

## ACKNOWLEDGMENTS

This work was supported by Ningbo Health Branding Subject Fund (PPXK2018-01), Medical and Health Science and Technology Plan Project of Zhejiang Province (grant no. 2018KY671 and 2019KY564), Major Social Development Special Foundation of Ningbo (grant no. 2017C510010). The content is solely the responsibility of the authors and does not necessarily represent the official views. The authors wish to gratefully acknowledge the patients who participated in this study to share their experiences. In addition, we express our heartfelt respect to all health care workers who are fighting against the COVID-19 epidemic.

